# Identification of dysregulated transcription factor activity in temporal lobe epilepsy

**DOI:** 10.1101/2025.04.24.25326321

**Authors:** Robert Zeibich, Terence J. O’Brien, Piero Perucca, Patrick Kwan, Alison Anderson

## Abstract

Approximately a third of patients with epilepsy continue to have uncontrolled seizures despite appropriate treatment with available antiseizure medications (i.e. drug-resistant epilepsy). There is emerging evidence that transcription factor (TF) activity is both dysregulated in epilepsy and modifiable by small molecule drugs, providing an opportunity for treatment innovation. Methods for identifying dysregulated TFs and their target genes are still in their nascent stage and the reproducibility of findings remains unclear. We aimed to determine concordance of findings, in terms of the TFs dysregulated, across different studies of drug-resistant epilepsy and to evaluate the performance of different methods for identifying dysregulated TFs. We used publicly available single nucleotide RNA-seq data to construct discovery and validation datasets comprising individuals with drug-resistant temporal lobe epilepsy and healthy controls. We found good concordance (83%, 105/126) between the pySCENIC (python implemented Single-cell Regulatory Network Inference and Clustering) and hdWGCNA (high-dimensional Weighted Gene Co-expression Network Analysis) methods. Cell-type specific concordance across the discovery and validation datasets was low (36% 137/377) and this could be attributed, in part, to differences in data quality. In contrast, we found strong concordance between TFs that met strict concordance criteria in the current study with those implicated in a tissue-level study in patients with drug-resistant epilepsies, with the overlap being higher for TLE-related modules relative to modules for other drug-resistant epilepsies [86% (32/37) vs. 21% (18/84), Fishers-exact test: 95% 7.40 to 85.5, p < 0.0001]. Most TFS identified had been reported as being associated with epilepsy in the overall literature (91%, 53/58). Our findings strengthen the hypothesis that TFs are key to the pathophysiology of drug-resistant epilepsy and could represent novel drug targets. We recommend that multiple methods be applied to optimise discovery.

## Introduction

Epilepsy is a neurological disorder characterised by an enduring predisposition to experience unprovoked epileptic seizures. It affects 3-4% of the global population over a lifetime, regardless of age, geography or ethnicity^1–4^. Seizures can be generalized, arising from both hemispheres of the brain, or focal, originating within networks limited to one hemisphere. While epilepsy can often be successfully treated with antiseizure medications (ASMs), approximately one-third of patients continue to have uncontrolled seizures despite appropriate ASM treatment^5^. Temporal lobe epilepsy (TLE) is the most common type of focal epilepsy in adults which is often resistant to ASMs. There is a pressing need to identify novel therapeutic targets to enable the development of therapies tailored to this form of epilepsy. Network-based methods applied to gene expression data are a powerful technique for elucidating mechanisms underlying pathology and identifying novel drug targets. One such method, the Causal Reasoning Analytical Framework for Target discovery (CRAFT) method^6^ was applied by Francois et al.^7^, to elucidate hallmarks of drug-resistant epilepsy including pathology-specific transcription factors (TFs) and their target genes.

Gene regulation is tissue- and cell-type specific and involves a complex interplay between epigenetic modifications and regulatory molecules including TFs and their target genes^8^. Given the cell type specificity, drugs that target TFs have the potential for highly specific disease modulation with fewer side effects^9,10^. Although TFs were originally considered to be “undruggable”, an increasing number of small molecules that directly or indirectly target TFs have entered clinical development^10^. The nuclear receptor family of TFs, which consists of 48 members, are the most druggable of TFs^9,11,12^. Regulatory units comprising a TF and its target genes are commonly termed gene regulatory networks (GRNs) or ‘regulons’ and identifying differential regulon activity between healthy and diseased states is a rapidly growing area of research. In neuropsychiatric disease, for instance, the Psychiatric Encyclopedia of DNA Elements (PsychENCODE) consortium brings together bulk and single-cell expression, chromatin and expression quantitative trait loci (eQTLs) data obtained from brain tissue and organoid model systems representing multiple disorders and healthy controls^13^, allowing the construction of cell type-specific gene regulatory networks, and disease-specific regulation of regulons^14^. In epilepsy, specific TFs have been implicated in epileptogenesis through diverse studies and are suggested to have potential as therapeutic targets^15^. These include *ZEB2*^16^, *CREB, REST/NRSF, NFKB, P53, P21, ARX,* and *KLF4*^15^. Transcriptional studies with human data are, however, limited and have mainly focussed on investigating differential expression^17,18^, the role of microRNA^19^, DNA methylation^20^ and validation of putative epilepsy genes^21^. To our knowledge, the study by Francois et al.^7^ was the first that comprehensively investigated gene regulatory networks in human brain tissue. This study compared tissue resected from patients with common subtypes of drug-resistant epilepsy, including a subset with TLE, with control post-mortem samples.

The search for disease-specific regulons is still in its nascent stage and the reproducibility of findings remains unclear. Regulon inference methods, which predict the interaction between TFs and their target genes, continue to evolve, including methods specific to single-cell sequencing which is important given the cell type specificity of gene regulatory mechanisms. Gold standards are yet to emerge, and the performance of any specific method is likely dependent on the quality and biological structure present in the data analysed. In this study, we aimed to determine concordance across discovery methods and datasets from different studies. We used publicly available single nucleotide RNA-sequencing (snRNA-seq) data to construct discovery and validation datasets comprising individuals with drug-resistant TLE and healthy controls. We applied two regulon inference methods to identify dysregulated TF activity at single-cell resolution. This allowed us to evaluate concordance across both datasets and methods to identify novel TLE-associated TFs and provide supporting evidence for TFs previously implicated in epilepsy.

## Results

### Sample characteristics

Unprocessed hippocampal snRNA-seq data from five individuals with drug-resistant TLE and six healthy controls were used as a discovery dataset, and processed hippocampal snRNA-seq data from four individuals with drug-resistant TLE and four healthy controls were used as a validation dataset (Table 1). All individuals with TLE underwent epilepsy surgery. Based on neuroimaging or histological analyses, none of the individuals with TLE in the discovery dataset had lesions, and in the validation dataset epileptogenic lesions were severe in two individuals, and two were categorised by the authors as ‘non/mild’. Available clinical characteristics for each dataset are shown in Table 2.

**Table 1:** Discovery and validation characteristics.

| TLE (GEO: GSE160189)<sup>22</sup><br>*Sequencing: Chromium 10x v2 and v3 (raw)*<br>*Samples: 5, Subsets: 10* |  |
| --- | --- |
| Sample ID | Age (Years) |
| 56_Batch2 (A56, P56) (Batch2) | 26 |
| 57_Batch3 (A57, P57) (Batch3) | 60 |
| 67_Batch5 (A67, P67) (Batch5) | 44 |
| 76_Batch6 (A76, P76) (Batch 6) | 24 |
| 82_Batch6 (A82, P82) (Batch 6) | 37 |

| Control (GEO: GSE186538)<sup>23</sup><br>*Sequencing: Chromium 10x v3 (raw)*<br>*Samples: 6, Subsets: 37* |  |
| --- | --- |
| Sample ID | Age (Years) |
| HSB231 (EC, SUB, DG, CA1, CA2-4) | 79 |
| HSB237 (EC, SUB, DG, CA1, CA2-4) | 51 |
| HSB628 (EC, SUB, DG, CA1, CA2-4) | 50 |
| HSB179 (DG1-7) | 48 |
| HSB181 (DG1-7) | 44 |
| HSB282 (DG1-8) | 48 |

**Validation**
| TLE (GEO: GSE216877)<sup>24</sup><br>*Sequencing: Chromium 10x v3 (processed)*<br>*Samples: 4, Subsets: 5* |  |
| --- | --- |
| Sample ID | Age (Years) |
| H-16-06-009 | 48 |
| H16-06-008 | 24 |
| H16-06-010 | 67 |
| H17-06-015 | 19 |

| Control (GEO: GSE185277)<sup>22</sup><br>*Sequencing: Drop-Seq (processed)*<br>*Samples: 4, Subsets: 5* |  |
| --- | --- |
| Sample ID | Age (Years) |
| Specimen_9 [S9] | 13 |
| Specimen_13 [S13] | 40 |
| Specimen_15 [S15] | 50.2 |
| Specimen_20 [S20] (1,2) | 89 |

DG= dentate gyrus, a=anterior hippocampus, P=posterior hippocampus, CA =cornu ammonis, EC=entorhinal corex, and SUB=subiculum.

**Table 2:** Patient characteristics.

| Discovery dataset<sup>23</sup> |  |  |  |  |  |
| --- | --- | --- | --- | --- | --- |
| Sample ID | Epilepsy duration (years) | Seizure frequency (per month) | Hemisphere | Neuropathology report summary | Medication |
| 56_Batch2 (A56, P56) | 11 | 0.5 | Left | No abnormalities | levetiracetam, lamotrigine |
| 57_Batch3 (A57, P57) | 37 | 3 | Left | No abnormalities | clonazepam, carbamazepine, pregabalin |
| 67_Batch5 (A67, P67) | 7 | 1.5 | Right | Reactive astrocytosis | lamotrigine, lorazepam |
| 76_Batch6 (A76, P76) | 15 | 1 | Left | No abnormalities | zonisamide, lamotrigine |
| 82_Batch6 (A82, P82) | 4 | 4 | Left | No abnormalities | lamotrigine, gabapentin |
| Validation dataset<sup>24</sup> |  |  |  |  |  |
| Sample ID | Seizure onset (years) | Seizure type (per month) | Hemisphere | Neuropath report summary | Medication |
| H16-06-008 | 17 | N/A | Left | None/mild HS | lamotrigine, divalproex, acetaminophen, fluticasone, doxycycline |
| H17-06-015 | 12 | 3-5 | Right | None/mild HS | depakote, clobazam, lamictal xr, topiramate, ascorbic acid, vitamin D, multivitamin |
| H16-06-009 | N/A | N/A | Left | Severe HS | levetiracetam, lamotrigine, lorazepam, zonisamide, phenytoin sodium extended, ergocalciferol, multivitamin |
| **H16-06-010** | N/A | 2-4 | Left | Severe HS | cyanocobalamin, levetiracetam, lacosamide, tiagabine, cholecalciferol. Ascorbic acid, aspirin, ascorbate calcium, multivitamine, docosahexanoic acid/EPA, lamotrigine, lisinopril, simvastatin, acetaminophen, setraline |

\*N/A: Information not available.

### Cell numbers and cell type annotation

Overall, fewer cells were detected in the validation dataset (n = 50,880) as compared to the discovery dataset (n = 284,646, Figure 1). Cell counts were more similar between groups in the validation dataset, comprising 25,451 TLE cells and 25,429 control cells, as compared to the discovery dataset, in which the TLE group had fewer cells (n = 83,545) than the control group (n = 201,101). When colouring the UMAP representation by group and sample, post-data harmonisation no visible batch effects were observed (Figure 3, B, C, E and F).

**Figure 1:**
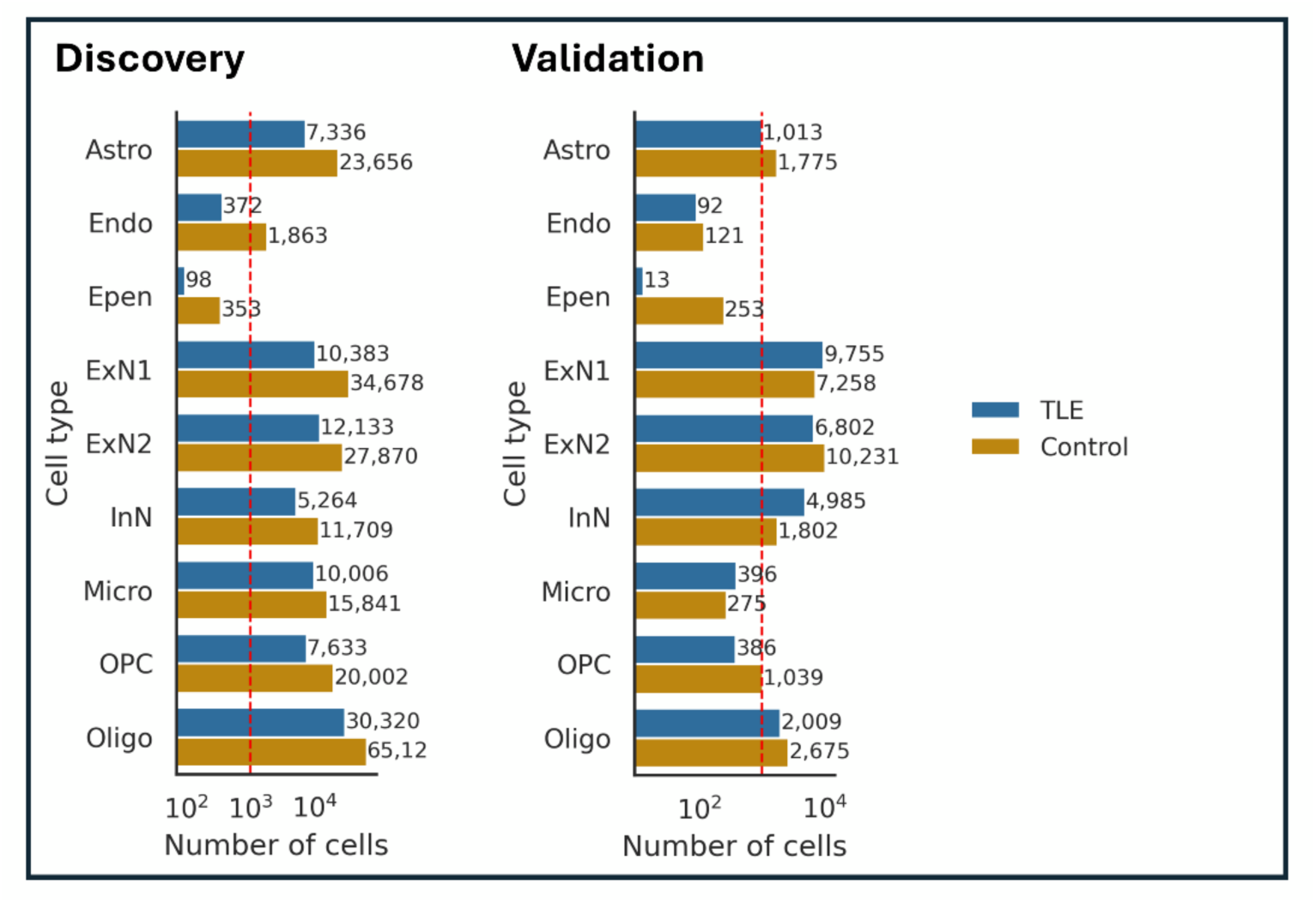
Cell type distributions. Total cell counts by cell type for the TLE and control group for the discovery and validation data. Abbreviations: astrocytes (Astro), endothelial cells (Endo), ependymal cells (Epen), excitatory neurons 1 (ExN1), excitatory neurons 2 (ExN2), inhibitory neurons (InN), microglia (Micro), oligodendrocytes precursors (OPC), oligodendrocytes (Oligo).

Following cell clustering expression levels of marker genes were used to assign cell type labels (Figure 2, Supplementary Figure S1-2). A total of 9 different cell clusters were identified in both the discovery and validation data: two groups of excitatory neurons (*SLC17A7* with *PPFIA, and TLL1*) and excitatory neurons 2 (*SLC17A7* with *PHACTR1*), inhibitory neurons (*GAD2*), oligodendrocytes (*PLP1, MBP*), oligodendrocyte progenitor cells (*TNR*), astrocytes (*AQP4*), microglia(*P2RY12*), endothelial (*COBLL1*), and ependymal cells (*CFAP54*) (Figure 3, A and D).

**Figure 2:**
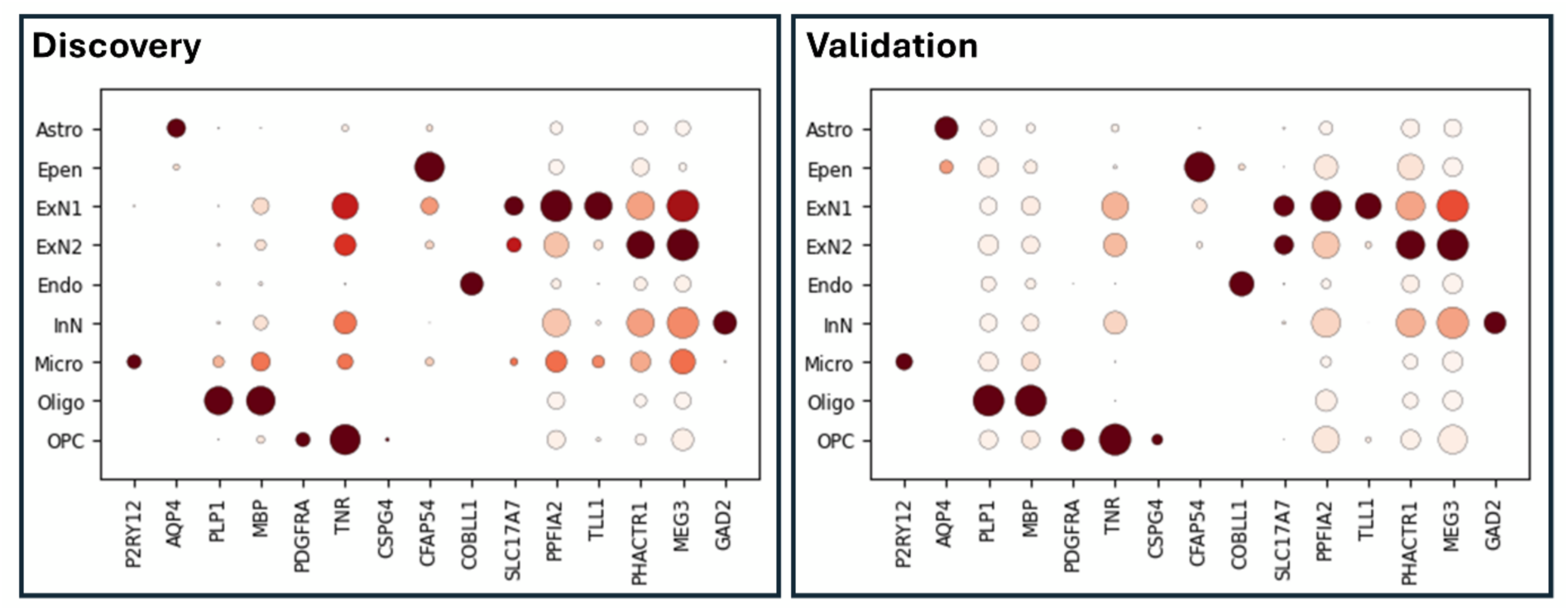
Marker gene Dot plots. Shows the expression levels and the fraction of cells (%) for a marker gene (x-axis) within each cell type (y-axis) for the discovery and validation data. The darker the colour the stronger the mean expression and the larger the circle the higher the fraction of cells expressing the marker in a cell cluster (%). Abbreviations: astrocytes (Astro), endothelial cells (Endo), ependymal cells (Epen), excitatory neurons 1 (ExN1), excitatory neurons 2 (ExN2), inhibitory neurons (InN), microglia (Micro), oligodendrocytes precursors (OPC), oligodendrocytes (Oligo).

**Figure 3:**
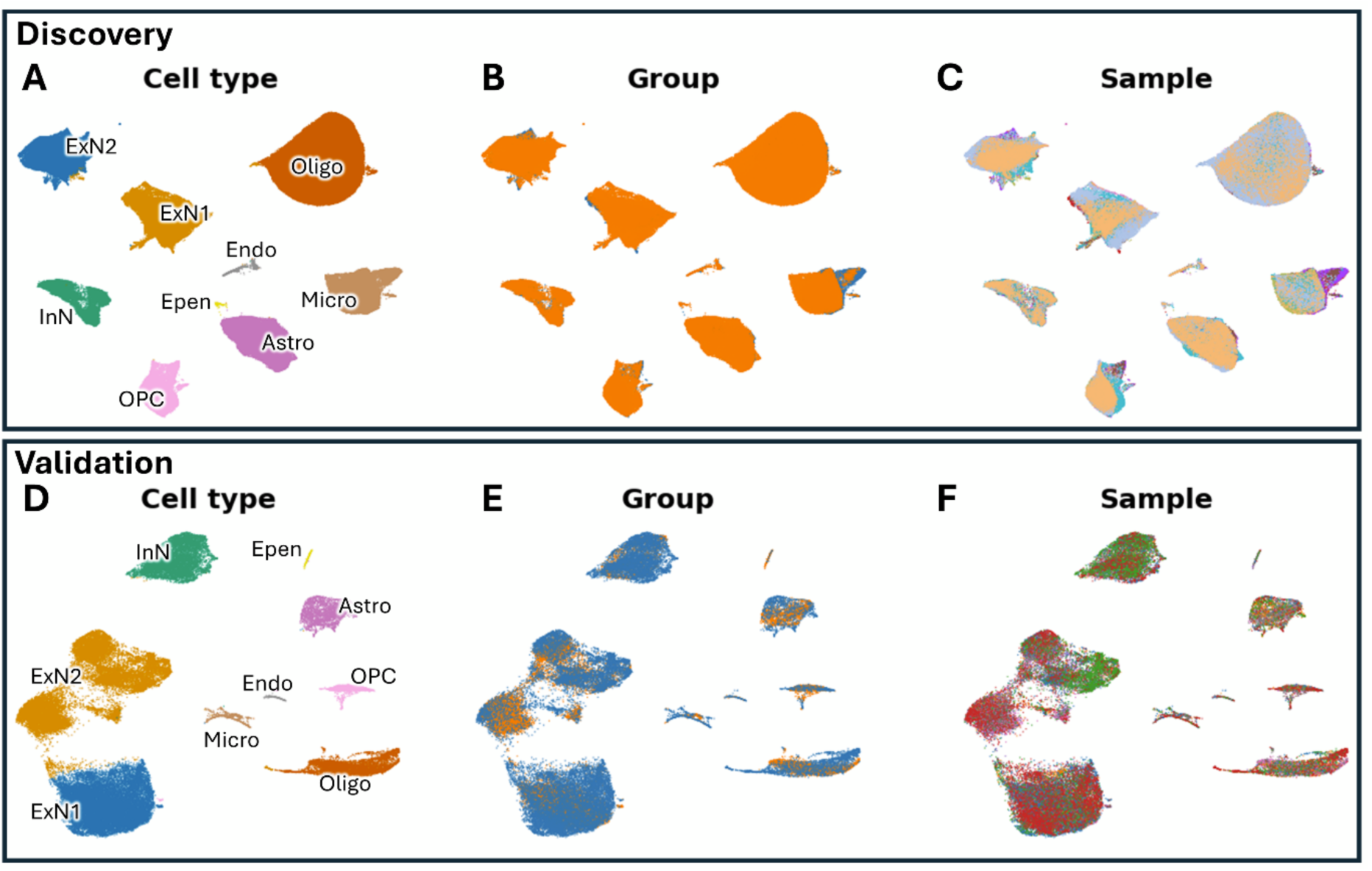
Batch corrected discovery and validation dataset. (A and D) UMAP representations coloured and annotated by cell type; (B and E) UMAP representation coloured by group (TLE =blue, Control = orange); (Cand F) UMAP representation coloured by sample. Abbreviations: astrocytes (Astro), endothelial cells (Endo), ependymal cells (Epen), excitatory neurons 1 (ExN1), excitatory neurons 2 (ExN2), inhibitory neurons (InN), microglia (Micro), oligodendrocytes precursors (OPC), oligodendrocytes (Oligo).

### Regulatory landscape

The regulatory landscape, generated from the regulon activity matrix, quantifies regulon activity across individual cells. When visualised as a t-SNE plot, with cells coloured by group, differences between groups are evident in both datasets (Figure 4), indicating that there is dysregulation between TLE and health control samples. In the discovery dataset, clusters are mostly similar for TLE and healthy controls, with differences being more evident within excitatory neurons 2, microglia, oligodendrocytes and astrocytes (Figure 4). In comparison, differences are more distinct across all clusters in the validation dataset and ependymal cells failed to form a separate cell cluster. This may be due to a higher level of technical artifacts in the latter, because, unlike the discovery dataset, only processed data were available, and different technologies were used to generate the TLE and healthy control datasets and different quality control and filtering criteria were likely applied.

**Figure 4:**
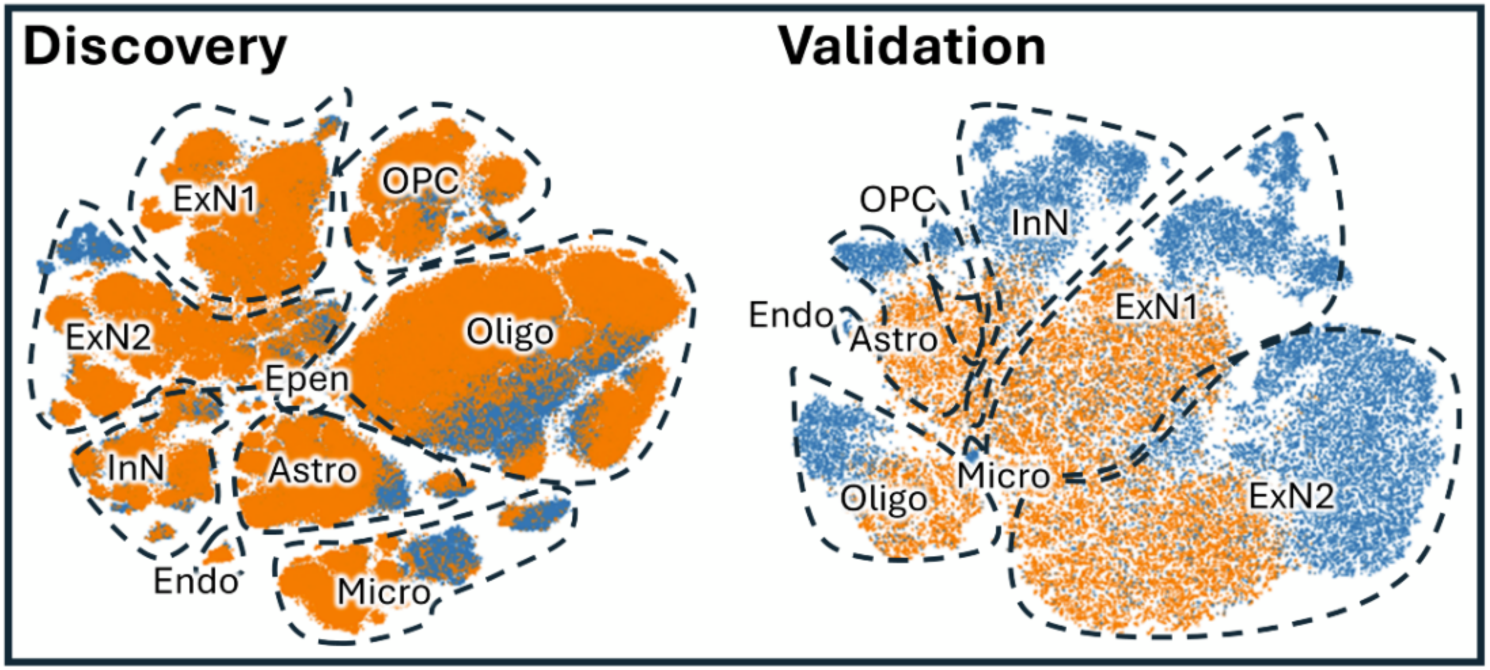
t-SNE representation by groups. Cells are coloured by group, TLE (blue) and healthy controls (orange) and clustered by cell type. Abbreviations: astrocytes (Astro), endothelial cells (Endo), ependymal cells (Epen), excitatory neurons 1 (ExN1), excitatory neurons 2 (ExN2), inhibitory neurons (InN), microglia (Micro), oligodendrocytes precursors (OPC), oligodendrocytes (Oligo).

### Dysregulated regulons

Dysregulation was determined by comparing cell level regulon activity scores and represents statistically significant differences in activity between TLE and healthy control cells for specific TFs, irrespective of differences in their associated target genes. Low cell counts prohibited the analysis of ependymal cells using the hdWGCNA method in both datasets and endothelial cells in the validation dataset (Figure 1). Overall, 983 statistically significantly dysregulated TF-regulons within cell type-specific clusters were identified across the discovery and the validation set. Concordance was assessed at three levels: (1) detection in the same cell type with the same direction (more active in either TLE or control cells) in both the discovery and validation datasets, (2) detection in the same cell type in both datasets irrespective of direction, and (3) detected in both datasets irrespective of both cell type and direction, as might be the case with tissue-level data. Only 7 % (72/983) met level 1 (Figure 5), and this increased to 20% (199/983) for level 2 and 36% (137/337) for level 3 (Supplementary Table S1). At level 3, large numbers of dysregulated regulons were found to be unique to the discovery dataset (n=137), or the validation dataset (n=103) (Supplementary Table S1). At level 1, dysregulation of TF activity was highest in astrocytes (n=14), followed by microglia (n=11), and inhibitory neurons (n=10, Figure 5).

**Figure 5:**
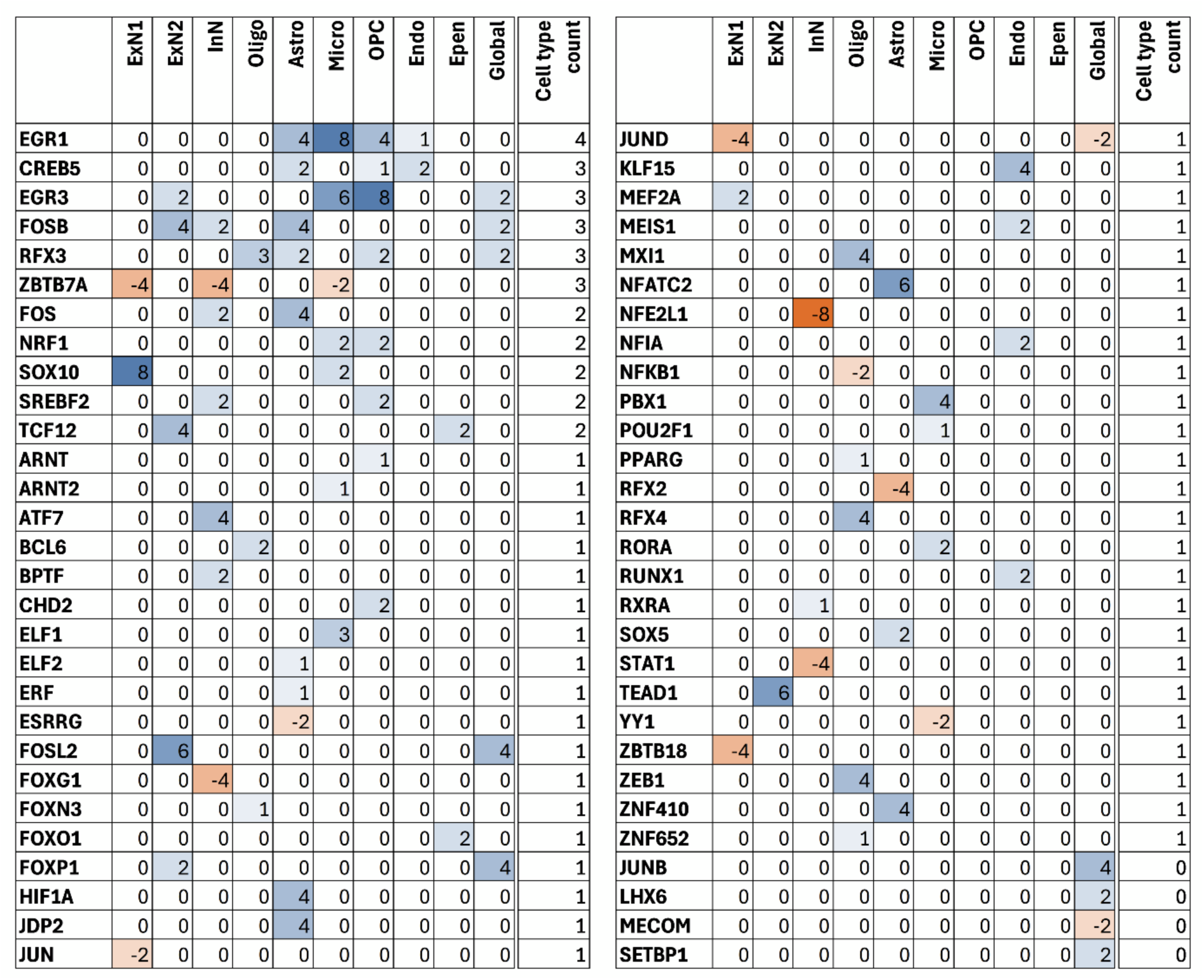
Dyregulated regulons at cell resolution. The positive number indicates higher regulon activity in epilepsy cells (n=59), while a negative number indicates higher regulon activity in control cells (n = 15). level of importance (pySCENIC: within top 15 highest mean absolute SHAP value =2, otherwise =1, hdWGCNA: within the top 15 most important =2, otherwise 1). The numbers were added at resolution across methods within each dataset and then multiplied at resolution across datasets. Higher values indicate stronger evidence with shade intensity increased (blue for epilepsy, orange for control). Abbreviations: astrocytes (Astro), endothelial cells (Endo), ependymal cells (Epen), excitatory neurons 1 (ExN1), excitatory neurons 2 (ExN2), inhibitory neurons (InN), microglia (Micro), oligodendrocytes precursors (OPC), oligodendrocytes (Oligo), across all cells (Global).

Across all cell types 41, dysregulated regulons were identified and of these 10 were in the same direction in both datasets and 6 (*EGR3, FOSB, FOSL2, FOXP1, JUND, RFX3*) had also been identified in specific cell types, while 4 (*JUNB, LHX6, MECOM, SETBP1*) were not (Figure 5, Global column). Twenty-seven were unique to the discovery dataset and four were unique to the validation dataset irrespective of directions (Supplementary Table S1).

Overall, across datasets concordance levels and methods applied, dysregulation predominantly involved higher activity in TLE (n=1,076 vs. 600) and five, within excitatory neurons 2, were active in both (determined by bimodal distribution on density plots), likely due to sub-clusters of different cell types within this specific cluster (see Supplementary Table S1 and Methods Cell annotation).

### TF with the strongest evidence of dysregulation

TFs driving dysregulated regulons that met the strictest concordance measures were ranked based on a high SHAP value for pySCNENIC and/or top-ranking by hdWGCNA (see methods for details). Seven obtained a score >= 6 (Figure 5), with six being active in TLE only (*EGR1, EGR3, SOX10, FOSL2, TEAD1* and *NFATC2*), and one, *NFE2L1*, more active in healthy controls. Among these, two were identified across all cell types and also within specific cell types: *EGR3* (excitatory neurons 2, microglia and oligodendrocytes precursor) and *FOSL2* (excitatory neurons 2, shown in Figure 6). Two were identified in multiple cell types: *EGR1* (astrocytes, microglia, oligodendrocytes precursor, and endothelial cells, shown in Figure 6), and *SOX10* (excitatory neurons 1, and microglia). The remaining 3 were detected in a single cell type: *TEAD1* (excitatory neurons 2), *NFATC2* (astrocytes) and *NFE2L1* (inhibitory neurons).

**Figure 6:**
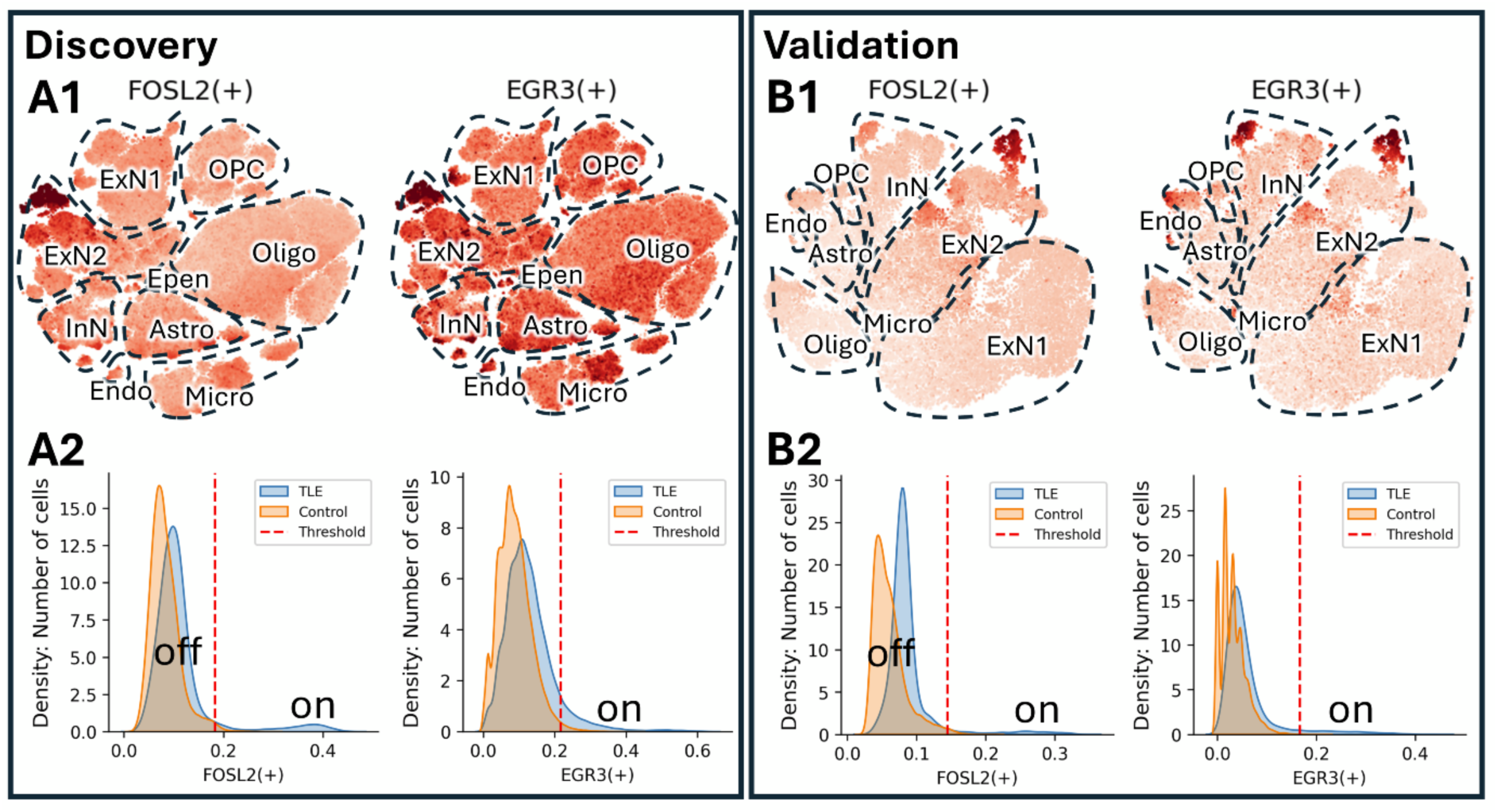
Regulon activity. (A1:B1) t-SNE representations: Cells are coloured based on the regulon activity (the darker the colour the higher the activity). (B1:B2) Density plots: Active (on) and inactive (off) cell populations in TLE and controls. Regulons are active in TLE cells only. Abbreviations: astrocytes (Astro), endothelial cells (Endo), ependymal cells (Epen), excitatory neurons 1 (ExN1), excitatory neurons 2 (ExN2), inhibitory neurons (InN), microglia (Micro), oligodendrocytes precursors (OPC), oligodendrocytes (Oligo), across all cells (Global).

### Concordance with literature

We looked for overlap between TFs implicated in our study and TFs implicated in gene expression modules associated with one or more of four common forms of drug-resistant epilepsies by Francois et al.^7^ Out of 121 modules, 37 were associated with TLE with HS (TLE-HS) and 84 with additional forms of drug-resistant epilepsy, comprising focal cortical dysplasia (FCD IIa and FCD IIb), and tubular sclerosis complex (TSC). We found that 73% (297/408) of TFs were present in one or more drug-resistant modules (Supplementary Table S2), including 86% (50/58) of those that met the strictest concordance measures, and all of the seven with the strongest evidence except for *FOSL2* (Supplementary Table S3). In our datasets, individuals with HS had been removed, as this condition was not the research focus of the initial studies. Nevertheless, the number of TFs impacting on modules associated with TLE-HS-related modules, either alone or in combination with modules associated with other types of refractory epilepsy, was significantly higher than for non-TLE-HS-related modules [86% (32/37) vs. 21% (18/84), Fishers-exact test: 95% 7.40 to 85.5, p < 0.0001] (Supplementary Table S3). One TLE-HS-related module (mTLE.20.o) reported by Francois et al.,^7^ to be the strongest delineator between TLE and healthy tissues, had the highest enrichment of TFs implicated in our study (n=7), comprising two among the highest ranked (*EGR1* and *EGR3*) and five dimers of the activator protein 1 (AP-1) complex (*FOS, FOSB, JUN, JUNB, JUND*), which is a master regulator of gene expression involved in response to various stimuli, including growth, differentiation, apoptosis, and immune response^25,26^.

Across the broader literature, 91% (53/58) of the TFs driving dysregulated regulons, that met the strongest measure of concordance, have been implicated in epilepsy including, three (*FOSL2, YY1, RFX3*) not reported in the study by Francois et al.^7^ One, *PPARG*^27^, was proposed as a potential treatment in epilepsy, based on a protective effect in a mouse model, and one, *NFKB1,* was among seven TFs suggested to be potential therapeutic targets by Sun et al.,^15^ five of which were identified in the discovery dataset only (*ARX, TP53, KLF4, REST, CREB1*), and one was not identified (*P21*). Five novel TFs were elucidated in this study, that to the best of our knowledge have not been associated with epilepsy (*JDP2, RXRA, RFX2, SREBF2, ZNF410*, Supplementary Table S3).

### Regulation of epilepsy-related genes

For dysregulated regulons meeting the strongest measure of concordance, we next investigated a possible role for epilepsy-related genes using the seizure-associated genes across species (SAGAS) database ^28^. We looked for overlap between human and rodent genes (n = 2,847) in this database and TFs driving dysregulated regulons (e.g. epilepsy genes that encode proteins that function as TFs), or gene targets within dysregulated regulons that were detected in both the discovery and validation datasets.

Approximately a third 34% (20/58) of TFs are epilepsy genes that encode proteins that function as TFs. A total of 269 target genes were epilepsy genes and these were regulated by 21 different TFs, of which 9 were also epilepsy related. The TFs that regulated the most epilepsy genes were: *CREB5* in oligodendrocytes precursor cells (n = 113), *FOSB* in inhibitory neurons (n = 38), *FOSL2* in excitatory neurons 2 (n = 27) and across all cell types (n = 30), *RFX3* in oligodendrocytes (n = 21), *FOSB* in excitatory neurons 2 (n = 18) and across all cell types (n = 20), *FOS* in inhibitory neurons (n = 17), *RFX3* in oligodendrocytes precursor cells (n = 13), and *NRF1* and *SREBF2* in oligodendrocytes precursor cells (n = 11, n = 10, Supplementary Table S4).

Epilepsy genes regulated by AP-1 dimers included *NPAS4, HOMER* and *BDNF* all of which contribute to epileptogenesis in a mouse model^29^, *ARC* which is significantly decreased in hippocampal tissue of rats with epilepsy and depression^30^ and three members of the *NR4A* orphan nuclear receptor family, *NR4A1-3,* that have been implicated in epilepsy through different investigative modalities: knockdown of *NR4A1* alleviated seizure severity in a mouse model^31^, de novo loss-of-function variants in *NR4A2* were identified in eight unrelated families with neurodevelopmental disorders and epilepsy^32^, and *NR4A3* was found to be upregulated following pilocarpine-induced seizures in a mouse epilepsy model^33^.

We also looked for overlap between target genes associated with epilepsy and the findings from Francois et al.,^7^ and found that 136 that were represented in TLE-HC modules and 11 (*FOSB, FOS, NR4A1-3, ARC, JUN, HASPA1A, KDM6B, TGIF1, PTGS2*) of these were within the mTLE.20.o module (Supplementary Table S5).

#### pySCENIC and hdWGCNA methods comparison

pySCENIC was more tolerant to cell clusters with low cell numbers and was able to detect dysregulated regulons in all 9 cell types whereas hdWGCNA could only detect within 8 in the discovery and 7 in the validation datasets. To compare performance, we considered only cell types common to both methods. For this subset, overall across both the discovery and validation dataset, pySCENIC identified 255 and hdWGCNA identified 179 significantly dysregulated regulons. The lower result for the latter could mostly be attributed to the tool being restricted to TFs listed in the JASPAR DB (n=1,227), 570 less than that evaluated by pySCNEIC (Supplementary Table S6). If only TFs available to both methods are considered then pySCENIC identified 155 while hdWGCNA identified 176 (with 105 in common), and both methods detected more dysregulated regulons in the discovery (pySCENIC=136, hdWGCNA=141) than in the validation dataset (pySCENIC=66, hdWGCNA=78), which we considered to potentially harbour more technical artefacts (Supplementary Table S7).

The tools evaluated, used different sources for TF lists. When considering TFs available to both methods, overall concordance was good (87% 105/126), but each tool had advantages and disadvantages. hdWGCNA identified more significant dysregulated regulons overall, while pySCENIC was more tolerant to small cell clusters. The authors of the tool claim it can identify cell types that are represented by only a few cells (e.g., two to six cells from microglia, astrocytes or interneurons)^34^. Using more than one detection method optimised detection. For instance, *CREB5* was only identified by pySCENIC, whereas *PPARG* was only captured by hdWGCNA (Supplementary Table S1). *PPARG* was found to be protective in a mouse model of epilepsy^27,35^ and out of all TFs driving dysregulated regulons, *CREB5* targeted the most epilepsy-associated genes and thus the identification of both is likely important. In terms of usability, hdWGCNA contains built-in functions to analyse dysregulated regulons thus requiring less programming than pySCENIC and the runtime was faster.

## Discussion

There is emerging evidence that TF activity is dysregulated in patients with drug-resistant TLE and other types of drug-resistant epilepsy. The development of drugs that target TF-driven mechanisms continues to evolve^10^, and provide opportunities for innovation in epilepsy treatment. The TF-driven regulatory landscape is complex, with cell type-specific nuance. Gold standard methods for detecting dysregulated activity are yet to emerge and the reproducibility of findings thus far remains unclear. To help address this knowledge gap, this study profiled regulon activity across two independent hippocampal snRNA-seq datasets from individuals with drug-resistant TLE and healthy controls using two different methods for detecting dysregulated TF activity.

At the strictest level of concordance, there was only 7% overlap in TF-driven regulons identified in the discovery and validation datasets, but this improved to 36% if cell type and direction (higher in TLE or higher in healthy controls) were not considered, as might be the case in tissue level analyses. Differences in capture technology and quality filtering criteria used in the studies likely contribute considerably to the poor concordance, and better results could be expected with more comparable datasets. The criteria for regulon detection by the software methods are strict and could be relaxed to increase yield, but potentially increasing false positives. We focussed our analyses on only TF-regulons that met our highest level of concordance criteria, and again this could have been relaxed to improve overall concordance. It was reassuring that many of the TFs implicated in this study, had been previously implicated in drug-resistant epilepsy^7^, and in particular, in a study specific to gene regulatory modules in brain tissue from patients with drug-resistant epilepsy^7^. We found the overlap with findings from the Francois et al.^7^ study to be strongest for modules specific to TLE-HS, despite differences between the studies in terms of the clinical characteristics (TLE with and without HS), granularity (tissue vs. single cell) and different approaches used to identify TF dysregulation. The tissue level study found enrichment of microglial cell-type markers while our analyses at the cellular level found most evidence for dysregulation within astrocytes, microglia and inhibitory neurons. Microglia have been shown to be increased in epileptogenic human and rat hippocampus^36^ and reactive astrocytes are associated with glial tissue, a histopathological characteristic of TLE^37^.

Components of AP-1, a dimeric TF complex, involving members of the Fos and Jun protein families^25^, were implicated in both studies. In a study that screened for functional impact of 476 regulatory variants in neurons and non-neurons from 616 human postmortem brains^38^, the top 8 neuronal TFs (enriched for variants that impact on their DNA binding affinity) were AP-1 components, including *FOSL2* implicated in this study. Depending on their composition, AP-1 complexes differentially bind to the DNA to drive cell type-specific changes in gene expression. AP-1 was also implicated in two studies by our group that investigated the teratogenicity of the ASM valproate ^39,40^ which involves a wide range of congential malformations and neurodevelopmental effects, and the ASM leviteracetam has been shown to reduce AP-1 activity in activated microglia in a mouse cell line via *FosL1*^41^.

Beyond epilepsy and psychiatric disease, dysregulation of AP-1 has been implicated in lymphomas^26^ and melanoma cells^42^. An important consideration here is the level of nuance and cell type specificity of TFs and the regulatory complexes they form. Analysis of the PsychENCODE data in neuropsychiatric disease, for instance, showed that the neural activity regulator and AP-1 component FOS have differential usage of enhancers and promoters across different psychiatric disorders^43^. This level of functional nuance is invaluable for understanding genetic predisposition, as genetic variation within specific enhancers and promoters could potentially delineate risk for certain disorders and explain comorbidities. At a broader level, Wang et al^44^ linked genetic variants associated with psychiatric diseases to a full gene regulatory network of the brain and when embedded into a deep-learning model these data gave a ∼6-fold improvement over polygenic risk scores for prediction of psychiatric phenotypes.

Cell type-specific nuance in TF complexes and their regulatory mechanisms is also important for the development of highly targeted therapeutics. In this study, *NFATC2* was among the top seven with strong evidence for dysregulation in TLE. This TF is a member of the NFAT family, associated with epilepsy^45^, that forms transcriptional complexes with AP-1 during an immune response. High throughput screening identified a small drug-like molecule that disrupts some but not all NFAT:AP-1 activity by binding in a sequence-selective manner to the DNA. This disruption of NFAT:AP-1:DNA interaction, provided improved specificity over existing immunosuppressive drugs and proof-of-concept that TF complexes on DNA are druggable^46^. Nuclear receptors are considered to be the most druggable family of TFs^9^. Several nuclear receptors were implicated in this study including the *NRA4* family and *PPARG* which are implicated in epilepsy and *RXRA* which we believe to be novel. Investigation of the activity of these regulators and their targets may expand therapeutic options beyond mechanisms more commonly targeted in epilepsy such as voltage-gated channels, γ-aminobutyric acid (GABA), neuronal excitation and synaptic function^47^.

For the discovery of TF dysregulation, we found that a tool can be limited by the source of TFs that was used in its design, and that the quality of the transcriptional data being analysed can influence performance. Given that methods have different advantages and disadvantages we recommend using at least two methods to optimise discovery. It should be noted that these methods are based on coexpression, and thus TF-gene target pairs can include two TFs that are part of a regulatory complex as well as non-TF gene targets, and as yet, these methods cannot delineate between these two scenarios.

A limitation of this study was that raw data were available for only one relevant dataset and processed data had to be used for validation. Preprocessing of raw single-cell data can have dramatic effects on downstream analyses, that can either improve discovery or limit potential by, for instance, removing small cell clusters that are biologically relevant. We believe that more comparable datasets and the availability of higher sample sizes would likely improve the level of concordance.

In conclusion, the concordance between our cell-level investigations and the tissue-level findings of Francois et al.^7^, and the literature in general, strengthens confidence in the biological relevance of dysregulation of TFs in TLE, and the opportunity for novel therapeutics that target these mechanisms. Our findings also underscore the value of applying multiple genomic bioinformatic methods to uncover dysregulated regulons.

## Methods

All snRNA analysis was performed using scanpy (v1.10.4).

### Pre-processing of discovery data

Publicly available raw hippocampal snRNA-seq data from five individuals with drug-resistant TLE, including five resected anterior and posterior human hippocampal tissue samples (GSE160189), along with raw snRNA data from six healthy individuals covering different hippocampal regions of the hippocampus (entorhinal cortex [EC], dentate gyrus [DG], subiculum [SUB], Cornu Ammonis one to four; GSE186538), were used for initial comparison. Samples from healthy individuals were postmortem, while samples from individuals with TLE were resected during surgery. The raw single-nuclei RNA (snRNA) reads were aligned to the genome build GRCh38 using simpleaf (v0.17.2) with --min-reads 1. QClus (v0.1.0)^48^, a pipeline specifically designed for snRNA seq data processing, was applied with default parameters to the aligned discovery data for quality control (QC) to remove ambient RNA, low-quality cells, including doublets using scrublet (v0.2.3).

### Pre-processing of validation data

At the time of analysis, additional raw snRNA-seq data for appropriate epilepsy cases was not publicly available for validation, thus processed hippocampal snRNA-seq data from four individuals with drug-resistant TLE (GSE216877) was used and compared with processed data from four healthy individuals with neurotypical hippocampi (GSE185277). Similar to the discovery dataset, samples from healthy individuals were postmortem, while samples from individuals with TLE were resected during surgery. These data had been aligned to genome build GRCh38. For the control data, cells expressing between 300 to 2,500 genes with a total count threshold above 500 and a mitochondrial gene percentage below 5% were retained. The drug-resistant TLE data had been pre-filtered. To check and improve data quality for both the drug-resistant TLE and healthy control data, doubletdetection (v4.1) and scrublet (v0.2.3) were applied to each data sample for doublet detection and each dataset was batch-corrected separately using Harmony, which included normalization, log transformation, identification of highly variable genes, scaling, and PCA computation. A nearest-neighbours graph was then constructed using 60 nearest neighbours based on the first 15 principal components, followed by UMAP embedding and clustering using the Leiden algorithm. Based on quality metric graphs and UMAP visualizations, Leiden clusters, predominantly composed of doublets, were removed and cells expressing between 500 and more than 12,500 genes were retained in the drug-resistant TLE dataset. In both datasets cells with a scrublet score below 0.15 were retained and cells with a complexity score between 0.8 and 0.99 were retained for the TLE data, and a stricter complexity score threshold of 0.9-0.99 was applied to the control data.

#### Discovery and validation dataset harmonisation and clustering

The datasets were batch-corrected and integrated separately using Harmony. The nearest-neighbours graph was computed with 50 nearest neighbours based on the first 17 components for the discovery data and 40 nearest neighbours based on the first 10 principal components for the validation data and UMAP embedding was generated with a minimum distance of 0.2 for both datasets. The Leiden algorithm was applied with a resolution of 0.2 for the discovery and 0.4 for the validation data. Three outlier Leiden clusters with fewer than 1,000 cells in fewer than three samples were removed from the discovery data.

### Cell annotation

To annotate Leiden clusters, we utilised marker genes reported to be specific to hippocampal cell types^49^ and the PandlaoDB^50^ resource as follows: *AQP4* identified astrocytes, *PLP1* and *MBP* oligodendrocytes, *PDGFRA*, *TNR and CSPG4* oligodendrocyte precursor cells, *P2RY12* microglia, *COBLL1* endothelial cells, *RBFOX2* and *SLC17A7* excitatory neurons, *GAD2* inhibitory neurons, and *CFAP54* ependymal cells. To validate the unique expression patterns of these marker genes in each cell type, we employed a dot plot and UMAP visualisations (Figure 2, Supplementary Figure S1-2). Two clusters were identified as excitatory neurons and were thus further classified into subtypes using gene expression data available in the ABC Atlas of cell types in the adult human brain^51^. *PPFIA2* and *TLL1* were highly expressed in the first cluster (excitatory neurons 1) in both datasets (Supplementary Table S8-9) and predominantly expressed in the hippocampus dentate gyrus (DG) cluster in the Atlas t-SNE plot. This is consistent with a t-SNE representation of tissue-level regulon activity, that we constructed, using information on tissue type from post-mortem controls in the discovery data, in which these markers were expressed in the dentate gyrus (Supplementary Figure S3). In contrast, highly expressed genes in the second cluster mapped to a broader range of cell types and regions including hippocampus CA1-3 (*MEG3*), CA4 (*MEG3*) eccentric medium spiny neurons (*PHACTR1, MEG3*), medium spiny neurons (*PHACTR1, MEG3*), amygdala excitatory (*MGE3*), and MGE interneurons (*MGE3, ROBO2*). This diversity of regions and neuronal cell types is in agreement with the t-SNE tissue regulon activity matrix representation of the discovery data (Supplementary Figure S3).

### Differential regulon analysis

#### pySCENIC

To identify regulons, we first applied the pySCENIC workflow, which uses the GENIE method to identify coexpression between TFs and genes (coexpression modules), and the RcisTarget method to filter those modules based on motif support within gene promoter regions for associated TFs and only modules with 10 or more target genes were used for downstream analyses. Non-normalised counts were used as input. The output is a matrix representing activity scores for all regulons in each cell. These scores were then used to determine which cells have higher subnetwork activity between TLE and healthy control groups. This software can be used to detect regulons within cell clusters and across all cells in a dataset and was applied in both contexts.

The discovery dataset contained 284,646 cells and evaluating the full set of TFs (1,797) with this many cells was computationally unfeasible. To identify and remove less informative TFs, we applied a screening method whereby all TFs were evaluated by iteratively (15 times) randomly selecting and analysing a subset of 40,000 cells per group. TFs that were found to be coexpressed with other genes in one or more of the iterations were selected for the regulon detection using all cells. This screening process was unnecessary for the smaller validation dataset.

#### hdWGCNA

We next applied high-definition Weighted Gene Coexpression Network Analysis (hdWGCNA), which is an extension of the standard WGCNA, designed to improve the robustness and interpretability of co-expression networks derived from sparse and high-dimensional data. hdWGCNA constructs metacells by aggregating small groups of similar cells within each defined group^52^, in our case TLE and healthy control, and cell type. Subsequently, eigengenes, summarizing gene expression profiles of co-expression modules, are computed, reducing the dimensionality. TF network analysis can then be applied to identify regulatory relationships between TFs and target genes using motif information from the JASPAR database (1,227 TFs). TF-driven regulons were derived using a strategy that selects the top TFs for each gene. For this method, the ependymal cell cluster was excluded from both datasets and the endothelial cell cluster from the validation dataset due to low cell counts.

### Identification of dysregulated regulons

For pySCENIC, regulons distinguishing the two groups were identified by applying the Mann-Whitney U test to the TLE and control regulon activity scores of each regulon from the regulon activity matrix, p < 0.05 was considered significant. To rank regulons, in terms of their capacity to delineate the two groups, we used the regulon activity scores with their corresponding group as a label, as input to an XGBoost model. To evaluate the contribution of each regulon to the model, we performed a Shapley Additive exPlanations (SHAP) analysis, whereas the mean absolute SHAP value was used as a measure of importance. While regulons with a high value can distinguish the two groups better, regulons with a value in the lowest 20% are less distinguishing and were therefore excluded. The top 15 regulons with the highest mean absolute SHAP value were annotated with a value of 2, while the remaining were annotated with a value of 1. To assess whether a regulon exhibits higher activity in TLE compared to healthy control cells (positive and negative activity direction, respectively), we calculated the median activity score for the TLE and control group for each regulon using the scores from the regulon activity matrix.

For hdWGCNA, statistically significant regulons distinguishing the two groups were identified using the in-built functions ‘FindDifferentialRegulons’ and ‘PlotDifferentialRegulons’, and given an order number using the function of the latter. Regulon with an order number below 15, indicating greater importance in distinguishing the two groups, were annotated with a value of 2 and the remaining were annotated with a value of 1. The degree of the activity direction was determined by the hdWGCNA metric avg_log2FC_positive. A value below zero indicates higher activity in TLE (positive direction) and a value above zero indicates higher activity in healthy controls (negative direction).

### Concordance between discovery and validation datasets

Concordance was assessed at three levels: (1) detection in the same cell type with the same direction (more active in either TLE or healthy control cells) in both the discovery and validation datasets, (2) detection in the same cell type irrespective of direction, and (3) detected in both datasets irrespective of both cell type and direction, as might be the case with tissue-level data. For TFs driving dysregulated regulons that met level 1 criteria, we looked for supporting evidence in the literature and in particular in the findings from Francois et al.^7^

### Regulation of epilepsy-related genes

Among dysregulated regulons that met level 1 criteria, we used pySCENIC to identify target genes in both the discovery and validation datasets. We then looked for TFs or target genes that were listed in the Seizure-Associated Genes Across Species (SAGAS) database. We identified TF-driven regulons that targeted the most epilepsy genes and epilepsy genes that were regulated by the most TFs.

### pySCENIC and hdWGCNA comparison

To compare the performance of both regulon detection methods, we compared the number of dysregulated regulons identified across the same cell types by both methods in each dataset, regardless of direction.

## Data Availability

All data produced are available online from the Gene Expression Omnibus repository.

https://www.ncbi.nlm.nih.gov/geo/query/acc.cgi?acc=GSE160189

https://www.ncbi.nlm.nih.gov/geo/query/acc.cgi?acc=GSE216877.

## Contributors

RZ processed, analysed, and interpreted the snRNA-seq data and drafted the manuscript. AA contributed to the data processing, analysis, and interpretation and contributed to the writing process. AA conceived and supervised the study. PK, TJO’B and PP were involved in supervision of RZ and critically revised the manuscript for intellectual content. All authors read and approved the final manuscript.

## Acknowledgement

This study was partly funded by the Australian National Health & Medical Research Council (NHMRC) (GNT1103979). PP P.P. is supported by an Emerging Leadership Investigator Grant from the from the National Health and Medical Research Council (APP2017651), The University of Melbourne, Monash University, and the Austin Medical Research Foundation. PK is supported by an NHMRC Investigator Grant (GNT2025849).

## Standard Protocol Approvals, Registrations, and Patient Consents

### Data availability

The data that support the findings of this study are available from the Gene Expression Omnibus repository: https://www.ncbi.nlm.nih.gov/geo/query/acc.cgi?acc=GSE160189 and https://www.ncbi.nlm.nih.gov/geo/query/acc.cgi?acc=GSE216877.

### Code availability

The workflow was implemented in Bash and Python and can be made available on request.

