## Supplementary Figures for "Identification of dysregulated transcription factor activity in temporal lobe epilepsy"

**Cell type annotation**

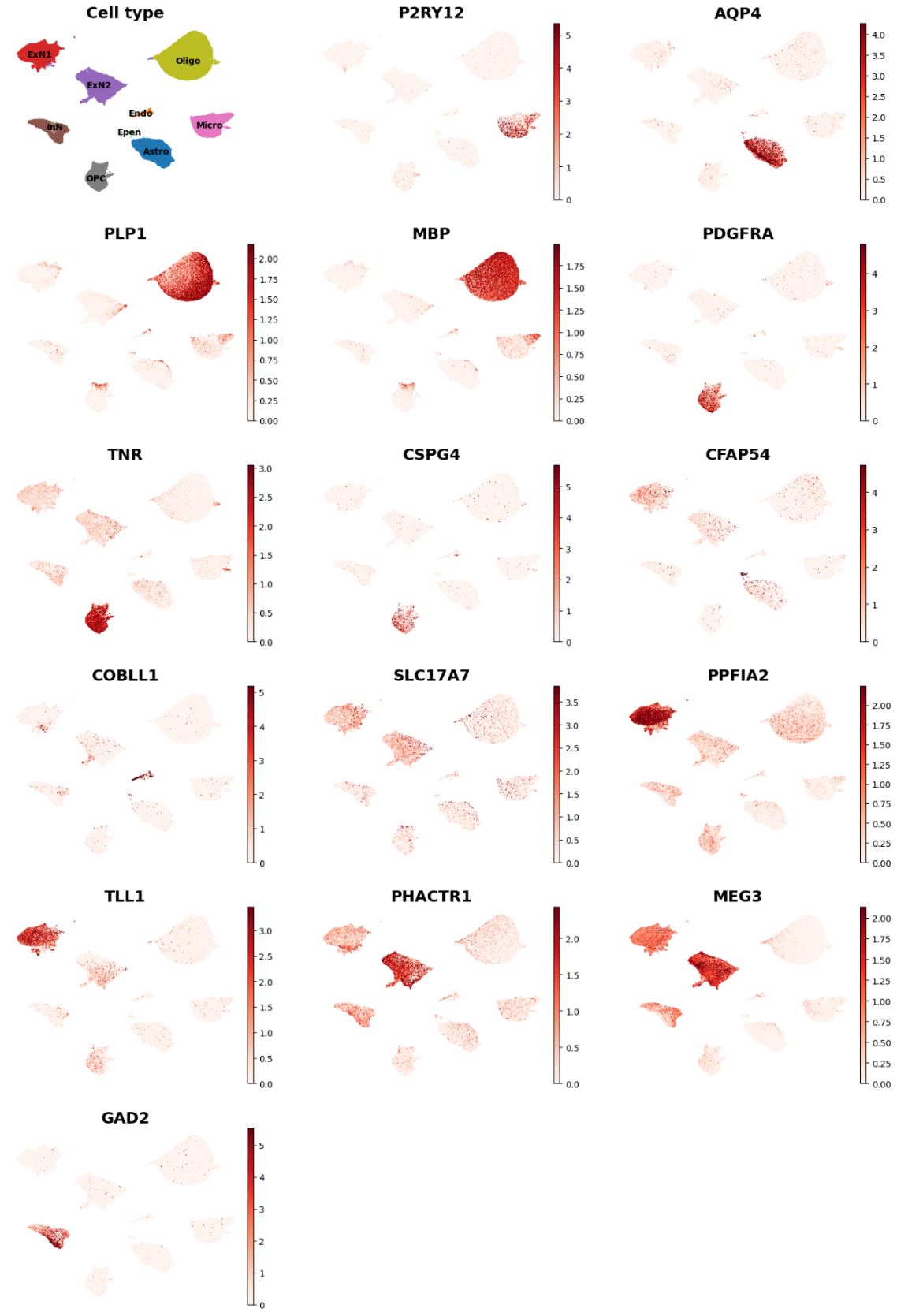

Figure 1: Marker gene expression discovery dataset. Batch-corrected UMAP plots of the the expression of marker genes for cell annotation. Abbreviations: astrocytes (Astro), endothelial cells (Endo), ependymal cells (Epen), excitatory neurons 1 (ExN1), excitatory neurons 2 (ExN2), inhibitory neurons (InN), microglia (Micro), oligodendrocytes precursors (OPC), oligodendrocytes (Oligo).

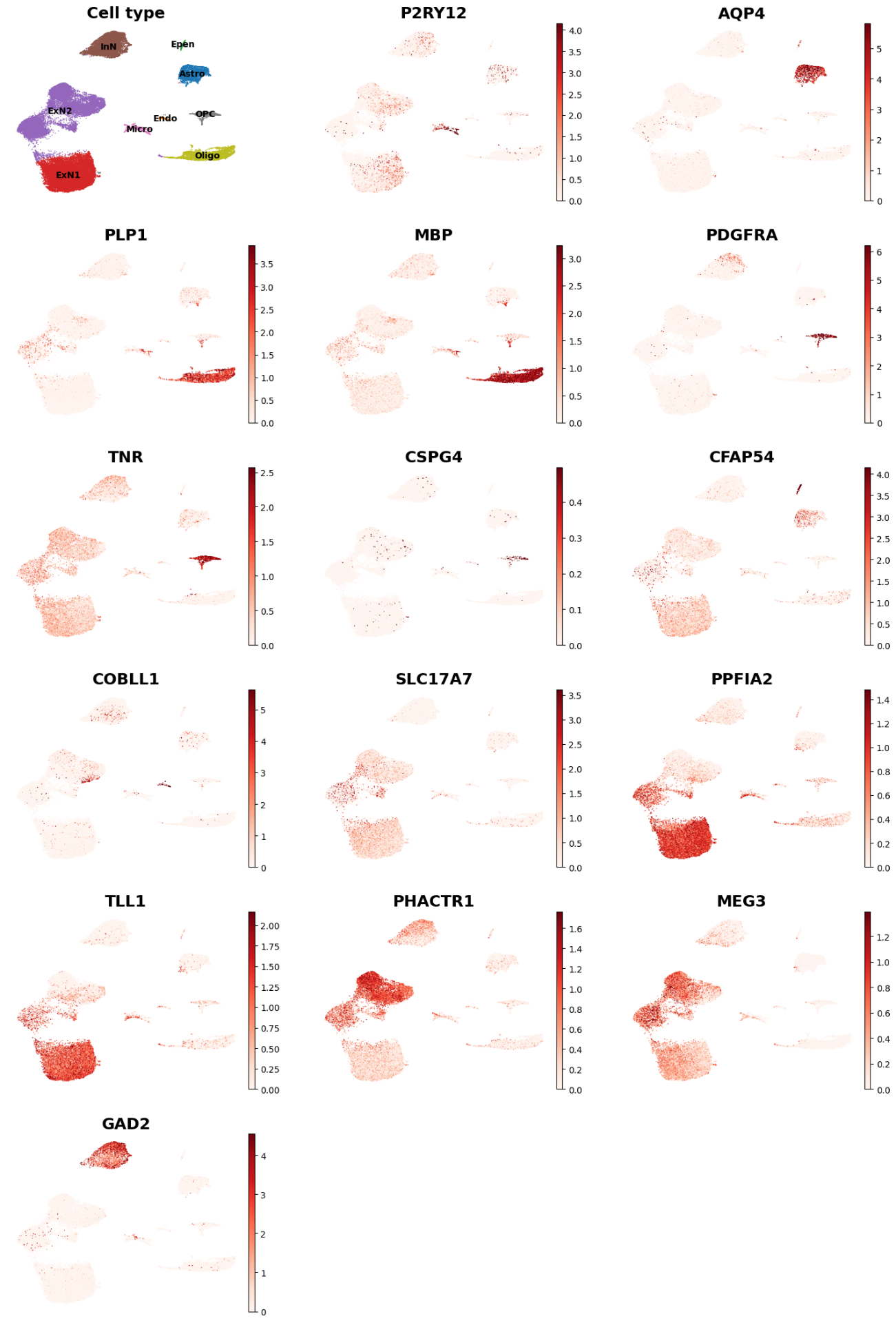

Figure 2: Marker gene expression validation dataset. Batch-corrected UMAP plots of marker genes for cell annotation. Abbreviations: astrocytes (Astro), endothelial cells (Endo), ependymal cells (Epen), excitatory neurons 1 (ExN1), excitatory neurons 2 (ExN2), inhibitory neurons (InN), microglia (Micro), oligodendrocytes precursors (OPC), oligodendrocytes (Oligo).

**Tissue representation**

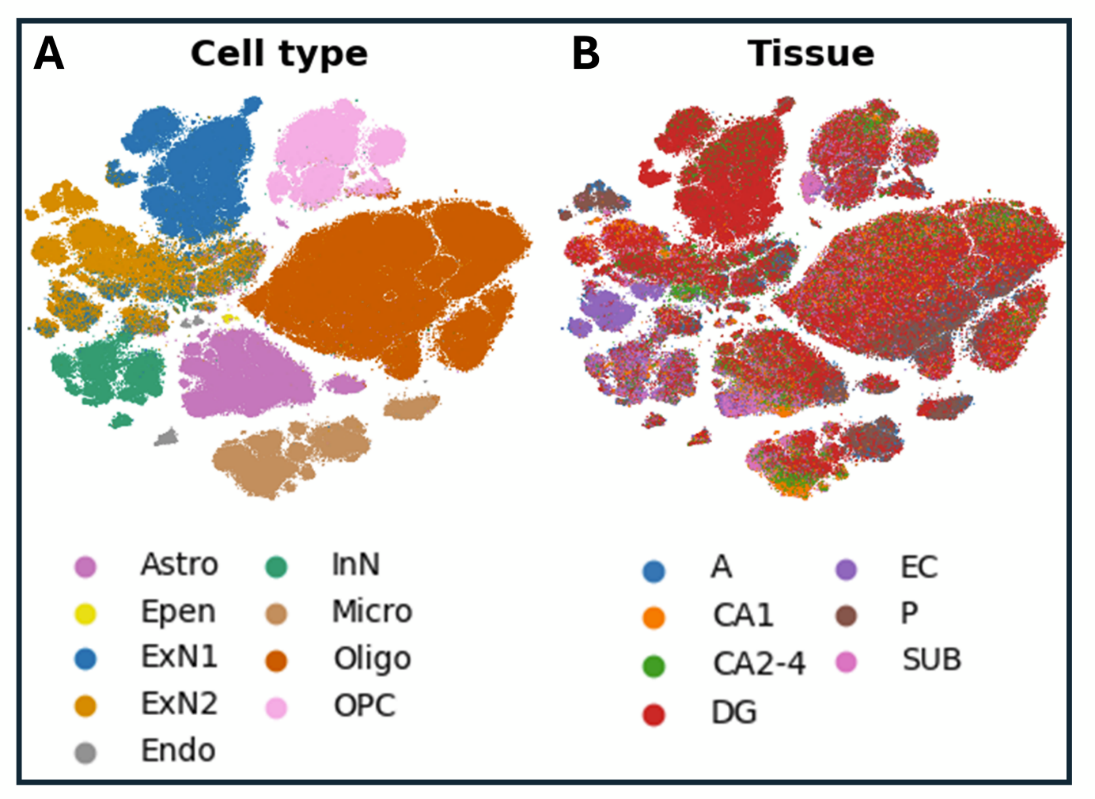

Figure 3 cell clusters coloured by tissue type. t-SNE representation of cells annotated with tissue informtion availble from the discovery dataset TLE (A and P) and post-motem samples, indicating that excitatory neurons 1 are enriched in the dentate gyrus (DG). Abbreviations cell types: astrocytes (Astro), endothelial cells (Endo), ependymal cells (Epen), excitatory neurons 1 (ExN1), excitatory neurons 2 (ExN2), inhibitory neurons (InN), microglia (Micro), oligodendrocytes precursors (OPC), oligodendrocytes (Oligo). Abbreviations tissue types: dentate gyrus (DG), anterior hippocampus (A), posterior hippocampas (P), cornu ammonis (CA1 and CA2-4), entorhinal corex (EC),subiculum (SUB).
